# A log-odds system for waning and boosting of COVID-19 vaccine effectiveness

**DOI:** 10.1101/2022.03.16.22272519

**Authors:** Tony Blakely, Joshua Szanyi, Tim Wilson, Nick Scott

## Abstract

Immunity to SARS-CoV-2 following vaccination wanes over time in a non-linear fashion, making modelling of likely population impacts of COVID-19 policy options challenging. We observed that it was possible to mathematize non-linear waning of vaccine effectiveness (VE) on the percentage scale as linear waning on the log-odds scale, and developed a random effects logistic regression equation based on UK Health Security Agency data to model VE against Omicron following two and three doses of a COVID-19 vaccine. VE on the odds scale reduced by 47% per month for symptomatic infection after two vaccine doses, lessening to 35% per month for hospitalisation. Waning on the odds scale after triple dose vaccines was 35% per month for symptomatic disease and 19% for hospitalisation. This log-odds system for estimating waning and boosting of COVID-19 VE provides a simple solution that may be used to parametrize SARS-CoV-2 immunity over time parsimoniously in epidemiological models.

## Introduction

Vaccines against severe acute respiratory syndrome coronavirus 2 (SARS-CoV-2) have repeatedly been shown to afford a high degree of protection from coronavirus disease 2019 (COVID-19) in the short term, particularly with regard to severe disease and death.^1-3^ Vaccine effectiveness (VE), however, is known to peak soon after receipt of a full vaccination course and wanes thereafter, an effect that has been demonstrated across multiple SARS-CoV-2 variants.^4-6^ This clearly poses significant obstacles to ongoing COVID-19 prevention and control efforts globally, but it also makes modelling of likely population impacts of COVID-19 policy options challenging.

There are now a plethora of published studies estimating VE over time following double and triple doses of multiple COVID-19 vaccines (e.g.^4,5,7^). In relation to the Omicron (B.1.1.529) SARS-CoV-2 variant, for example, protection against symptomatic disease following a primary course of the BNT162b2 (Comirnaty, Pfizer-BioNTech) vaccine has been shown to fall from 65.5% at 2-4 weeks following receipt of a second dose to 8.8% at ≥25 weeks.^4^ After a booster dose of BNT162b2, protection increases to 67.2% at 2-4 weeks which then decreases to 45.7% by 10 weeks.^4^ Synthesising these observations for use within simulation models, however, is not immediately straightforward.

As VE is bound between 0% and 100%, a similarly bounded log-odds system may provide a parsimonious solution to this problem. When plotted on a percentage scale, the gradient of vaccine-derived immunity waning often tends to be flatter soon after completion of a vaccine course and then steepens over time, before flattening out again as it asymptotes to 0%. This is consistent with linear decline in the log-odds when plotted back onto a percentage scale. A linear decrease in the log-odds is equivalent to a constant odds ratio (OR) applied to the VE odds every month. For example, consider an initial VE of 90%, which is 9 on the odds scale (90%/10%). Assuming an OR of waning over time of 0.6, the VE odds in months 2 to 6 would be 0.6 × 9 = 5.40; 0.6 × 5.40 = 3.24; 1.94; 1.17; and 0.7. Converted back to the percentage scale for months 1 to 6 this would be 90%; 84%; 76%; 66%; 54%; and 41%. Note the increasing monthly percentage point reduction, as observed in the many studies of VE over time. Note also that as VE falls below 50%, the steepening of the decline (on the percentage scale) begins to slow: extending the above series for months 7 to 10 gives VE on the percentage scale of 30%, 20%, 13% and 8%, respectively.

These properties, especially that we could mathematize non-linear waning on the percentage scale as simple linear waning on the log-odds scale, prompted us to test our schema on UK Health Security Agency (UKHSA) data for VE against Omicron for symptomatic illness and hospitalization.^8^

## Methods

Point estimates of VE against symptomatic disease and hospitalisation from infection with the Omicron variant on the percentage scale, and their corresponding upper and lower confidence limits, were extracted from tables (at ^4^) and visually from graphs for latest data (from ^8^) for all available ‘sub-studies’ (each different combination of vaccine course and outcome). The log-odds for all data points were then calculated, as well as the inverse variance of each observation (the variance being the difference between the 95^th^ and 2.5^th^ percentile limits on the log odds scale, divided by 3.92, then squared).

The following random effects logistic regression (with separate class for each ‘sub-study’) was fitted, excluding observations within 2 weeks since last vaccine dose (given peak immunity would still have been developing), weighted by the inverse variance:

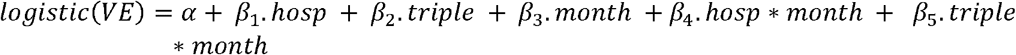

where *hosp* is a dummy variable for hospitalisation versus symptomatic illness; *triple* is a dummy variable for triple versus double vaccine course; *month* represents months since last vaccine dose minus 0.5 (i.e. ‘centered’ on two weeks post the last vaccine dose to aid interpreting model coefficients); *hosp*month* is an interaction term of hospitalisation and month; and *triple*month* is an interaction of triple and month. We also fitted a model with a three-way interaction of *hosp*triple*month*, but the coefficient was non-significant and the difference in deviance statistic with the above model was trivial and non-significant. As such, this term was not retained in the final model.

## Results

The coefficients from the random effects logistic regression model are shown in Table 1. Converting the intercept to the percentage scale gives a VE of 67.7% (95% confidence interval [CI] 62.1% to 72.8%) for the reference case of VE against symptomatic Omicron infection, two weeks post second dose. The OR for VE against hospitalisation compared to VE against symptomatic disease is 3.11 (95% CI 1.87 to 5.16), and for triple compared to double dose is 1.10 (95% CI 0.83 to 1.45). VE waning has an OR of 0.53 for symptomatic infection after two doses, or a 47% reduction in the VE odds per month. For hospitalisation the VE odds reduces by 35% per month (i.e. 1 minus the OR of 0.65, where 0.65 = 0.53 × 1.24). After triple dose vaccination, the VE odds for symptomatic disease also decreases by 35% per month (1 minus 0.53 × 1.23), and the VE odds for hospitalisation decreases by 19% per month (1 minus 0.53 × 1.24 × 1.23).

**Table 1:**
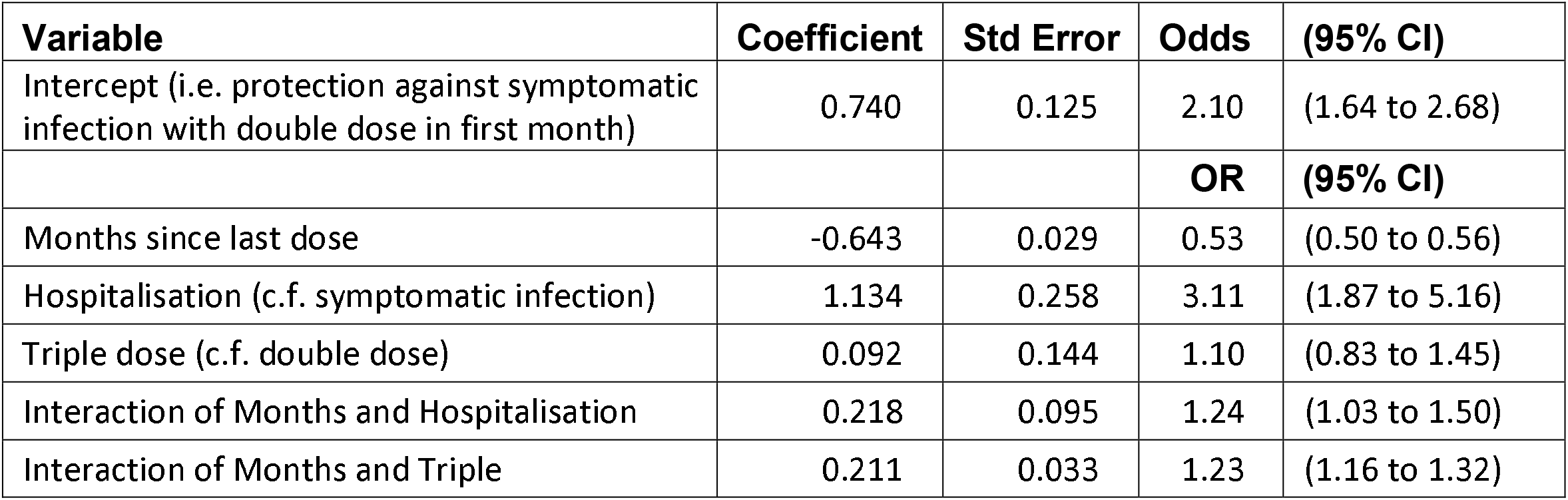
Logistic regression coefficients and odds ratios for vaccine efficacy against the Omicron SARS-CoV-2 variant

Figure 1 presents the predicted VE from the regression model converted to the percentage scale with the data points used to develop the regressions overlaid.

**Figure 1:**
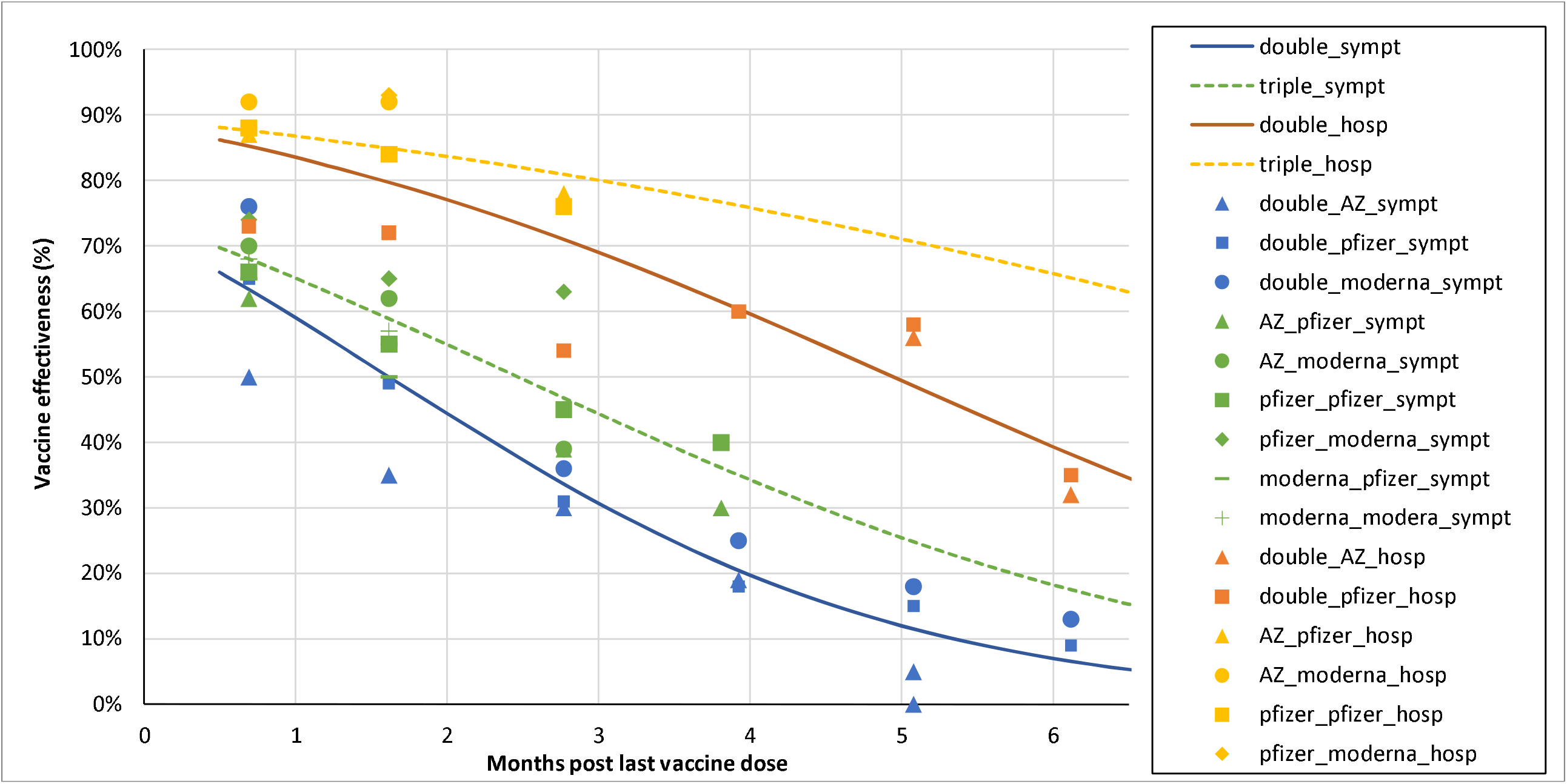
Predicted vaccine effectiveness against the Omicron SARS-CoV-2 variant using a log-odds system, overlaid with observed data points used to fit model. Confidence bands about the predictions are shown in the Supplementary Figure 1

## Discussion

Waning of immunity following COVID-19 vaccination presents a challenge when attempting to model likely population impacts of COVID-19 policy options. As more has been learnt about the nature of immunity to SARS-CoV-2, it has been necessary for models to move away from assuming constant VE to incorporating some type of waning function. These functions can vary in complexity from step functions to linear declines or more explicit relationships between neutralizing antibodies and protection against infection, symptomatic disease, hospitalisation, and death. Our analysis shows that a single simplifying assumption that waning immunity takes a log-odds functional form can explain reasonably well the observed profiles of protection across doses and outcomes, which may considerably assist in the parametrization of epidemiological COVID-19 models. Overlaying the data points used to develop the regression model onto the predicted VE provides a reasonable fit.

Importantly, the log-odds system we have developed can be easily extended. First, it can be expanded to include more observations from the UKHSA data series as they are generated. Second, data from additional studies in other settings could be included. For example, data from Brazil could be incorporated into the model,^9^ noting that in countries with a less effective primary course vaccine, the VE boost with a third dose mRNA vaccine (i.e. the OR of a triple compared with a double dose) may be larger than in the UK.

Third, the system can be reconfigured to include additional covariates. For example, there is good evidence of lesser VE among older people following vaccination, particularly for symptomatic disease.^5^ Even if age-related data are not available for the regression model (as was the case with the UKHSA data we used to develop our model), one can still ‘add’ an extra variable to separate VE estimates by age – ensuring the weighted average by age of VE still returns that obtained in the starting model (e.g. by adjusting the intercept).

Fourth, importantly for modelling, this mathematical system can be extended to include ‘known unknowns’. For example, there may be a desire in future to undertake scenario modelling of next-generation vaccines or new variants of concern. If plausible scenarios regarding the ‘boost’ in VE compared to triple dose of current-generation vaccines or relative immune escape are available, it is straightforward to extend these equations for use in simulation modelling of these scenarios.

Ultimately, for population-level mathematical models to be useful they need to include assumptions about the functional form of waning immunity, and how it differs by outcome (for example, symptomatic infection compared to hospitalisation) and the number of vaccine doses received. This log-odds system for estimating waning and boosting of COVID-19 VE provides a simple to apply solution that may be used to more accurately approximate SARS-CoV-2 immunity over time when parametrizing epidemiological COVID-19 models.

## Data Availability

All data produced in the present study are available upon reasonable request to the authors.

---

**Supplementary Figure 1:**
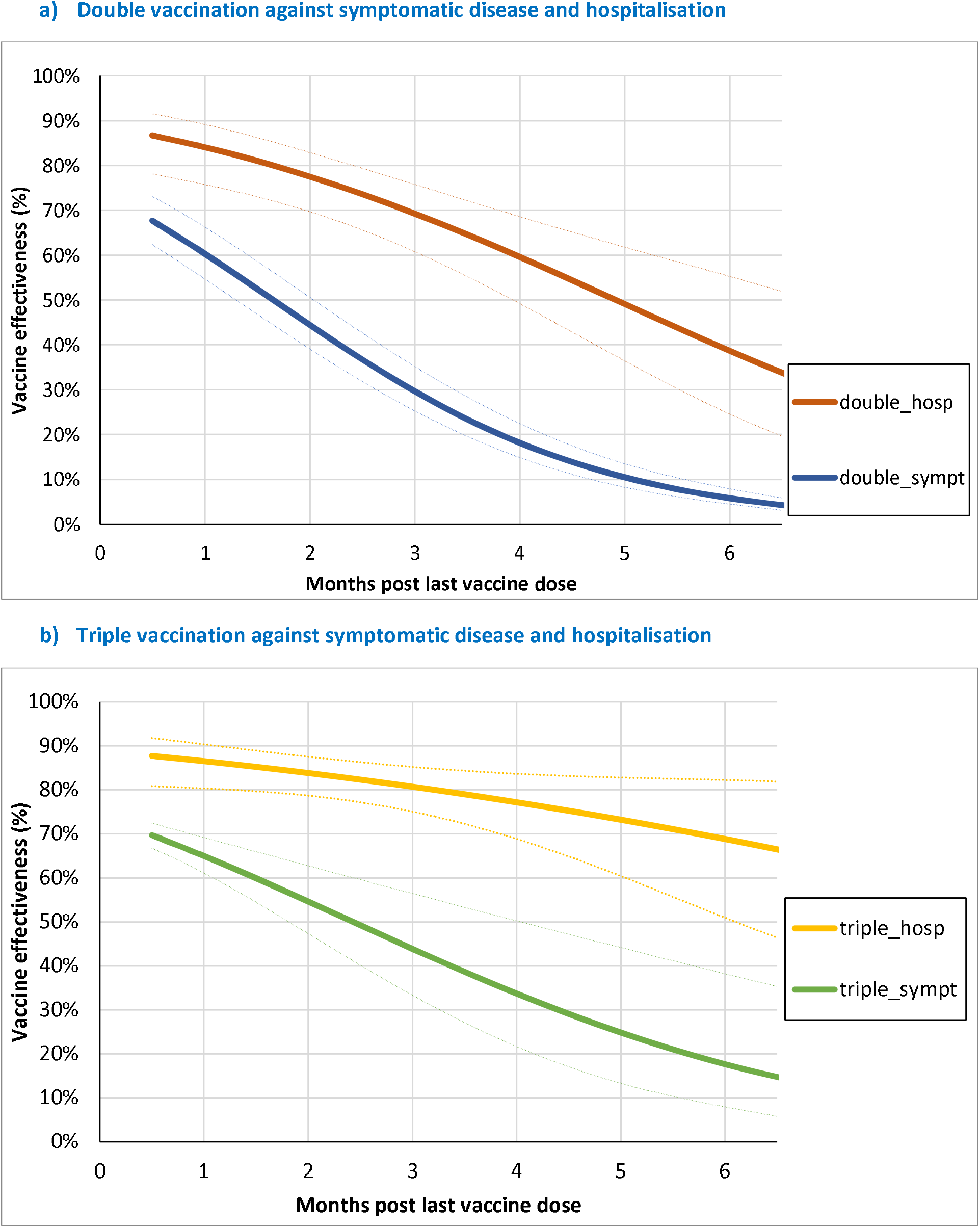
95% confidence bands about predicted vaccine effectiveness against the Omicron SARS-CoV-2 variant using a log-odds system.

## Notes

### Competing Interest Statement

The authors have declared no competing interest.

### Funding Statement

This study did not receive any funding.

### Author Declarations

This study involves only openly available human data, which can be obtained from: https://www.gov.uk/government/publications/covid-19-vaccine-weekly-surveillance-reports

